# An examination of COVID-19 mitigation efficiency among 23 countries

**DOI:** 10.1101/2020.08.23.20180554

**Authors:** Yi-Tui Chen, Shih-Heng Yu, Emily Chia-Yu Su

## Abstract

The purpose of this paper is to compare the relative mitigation efficiency of COVID-19 transmission among 23 selected countries, including 19 countries in the G20, two heavily infected countries (Iran and Spain), and two highly populous countries (Pakistan and Nigeria). This paper evaluated the mitigation efficiency for each country at each stage by using data envelopment analysis (DEA) tools and analyzed changes in mitigation efficiency across stages. Pearson correlation tests were conducted between each change to examine the impact of efficiency ranks in the previous stage on subsequent stages. An indicator was developed to judge epidemic stability and was applied to practical cases involving lifting travel restrictions and restarting the economy in some countries.

The results showed that Korea and Australia performed with the highest efficiency in preventing the diffusion of COVID-19 for the whole period covering 120 days since the first confirmed case, while the USA ranked at the bottom. China, Japan, Korea and Australia were judged to have recovered from the attack of COVID-19 due to higher epidemic stability.

## Introduction

The COVID-19 pandemic has been raging across the world since the beginning of 2020, resulting in a substantial death toll. At the present time (2020/07/12), the World Health Organization (WHO) has indicated that more than 216 countries, areas or territories have been found to have 12,552,765 confirmed cases associated with 561,617 deaths [1]. The number of daily, newly confirmed cases in some countries has fallen to single or double digits, but in some other countries, it has not reached its peak and continues to increase. This shows that the response strategies adopted in each country may have different effects on the mitigation of COVID-19 transmission.

In this paper, we attempt to measure the relative efficiency in preventing the spread of COVID-19 by using the data envelopment analysis (DEA) technique. In practice, the DEA technique has been widely used in various applications, including health industries (e.g., Yaya *et al*. [2]; Cavalieri *et al*. [3]), energy sectors (e.g., Balitskiy *et al*. [4]; Cruz *et al*. [5]; Vazhayil and Balasubramanian [6]), cement industries (e.g., Oggioni *et al*. [7]), agricultural production (e.g., Mousavi-Avval *et al*. [8]; Mohammadi *et al*. [9]), and manufacturing sectors (e.g., Egilmez *et al*. [10]), and DEA has proven to be an effective approach in identifying the best practice frontiers.

In the field of medical services, DEA has also been widely used to measure the efficiency of hospitals in association with patient visits, surgeries and discharges. For example, Khushalani and Ozcan [11] employed a dynamic network DEA to examine the efficiency of producing quality in hospitals and found that urban and teaching hospitals were less likely to improve quality production efficiency. Deily and McKay [12] used efficiency scores obtained from a DEA analysis as explanatory variables to determine hospital efficiency. In other fields, Oggioni *et al.* [7] employed DEA to analyze efficiency by using energy as an input and one desired output accompanied by undesired outputs (CO2 emissions). Mousavi-Avval *et al*. [8] and Mohammadi *et al*.

[9] applied the DEA technique to measure the efficiency of agricultural production to identify wasteful energy. Vazhayil and Balasubramanian [6] showed that the weight-restricted stochastic DEA method was appropriate to optimize power sector strategies.

To compare the mitigation efficiency among countries on a fair basis, the time period for each stage was calculated from the date of the first confirmed case in each country. The whole period covers 120 days from the first confirmed case and was divided into 6 stages. In addition to the measurement of overall efficiency covering 120 days, the efficiency at each stage was also evaluated. The trends in efficiency ranks across stages for each country were analyzed. Eventually, an indicator for epidemic stability was developed to judge the status of epidemic stability for each country.

## Research methods

To compare the relative prevention efficiency to reduce the spread of COVID-19, a total of 23 countries were selected, including 19 countries in the G20 and the other four representative countries, as listed in Table 1. The reasons for the selection of Iran and Spain was due to their high levels of confirmed cases and deaths. Pakistan and Nigeria were chosen due to their large populations, which reached 212.2 million and 195.9 million, respectively, at the end of 2018 [13].

The WHO [1] has divides the stages of transmission into ( 1) no cases reported or observed ( Stage 0); ( 2) imported cases ( Stage 1); ( 3) localized community transmission ( Stage 2); and ( 4) large-scale community transmission ( Stage 3). As the date of the first confirmed case varied across countries, the period of each stage was not based on the same date among these countries but was calculated from the first confirmed case in each country. The date of the first confirmed case was identified based on the daily situation report released by the WHO [1] starting on 21 January 2020. Among the 23 counties selected, China, Japan, and Korea reported having confirmed cases of COVID-19 before 21 January 2020. The information released from the WHO [1] demonstrated that some cases of pneumonia of unknown etiology were detected in Wuhan City, Hubei Province, China, on 31 December 2019. On 7 January 2020, a new type of coronavirus was isolated and identified. Thus, the first case in China may be considered to have occurred at the end of 2019. According to the WHO [1], the first confirmed cases of COVID-19 in Japan and Korea were reported on 15 and 20 January 2020, respectively.

The overall efficiency was compared based on the whole period covering 120 days since the first confirmed case for each country. This paper separated the development process of COVID-19 spread into 6 stages. Stage 1 covers the first 30 days after the first confirmed case in each country. Each stage from Stage 2 to Stage 6 covered 15 days each. days after the first confirmed case in each country. Each stage from Stage 2 to Stage 6 covered 15 days each. In summary, the starting and ending dates of each stage for each country are listed in Table 1.

**Table 1:** The starting and ending dates of each stage for each country.

|  | Stage 1 | Stage 2 | Stage 3 | Stage 4 | Stage 5 | Stage 6 |
| --- | --- | --- | --- | --- | --- | --- |
| China | 2019/12/31-2020/1/30 | 2020/1/31-02/14 | 02/15-02/29 | 02/30-03/15 | 03/16-03/30 | 03/31-04/14 |
| Japan | 01/15-02/14 | 02/15-02/29 | 03/01-03/15 | 03/16-03/30 | 03/31-04/14 | 04/15-04/29 |
| Korea | 01/20-02/19 | 02/20-03/05 | 03/06-03/20 | 03/21-04/04 | 04/05-04/19 | 04/20-05/04 |
| USA | 01/23-02/22 | 02/23-03/08 | 03/09-03/23 | 03/24-04/07 | 04/08-04/22 | 04/23-05/07 |
| Australia,<br>France | 01/25-02/<br>24 | 02/25-03/<br>10 | 03/11-03/<br>25 | 03/26-04/<br>09 | 04/10-04/<br>24 | 04/25-05/<br>09 |
| Canada | 01/27-02/<br>26 | 02/27-03/<br>12 | 03/13-03/<br>27 | 03/28-04/<br>11 | 04/12-04/<br>26 | 04/27-05/<br>11 |
| Germany | 01/28-02/<br>27 | 02/28-03/<br>13 | 03/14-03/<br>28 | 03/29-04/<br>12 | 04/13-04/<br>27 | 04/28-05/<br>12 |
| India | 01/30-02/<br>29 | 03/01-03/<br>15 | 03/16-03/<br>30 | 03/31-04/<br>14 | 04/15-04/<br>29 | 04/30-05/<br>14 |
| Italy | 01/31-03/<br>01 | 03/02-03/<br>16 | 03/17-03/<br>31 | 04/01-04/<br>15 | 04/16-04/<br>30 | 05/01-05/<br>15 |
| Russia,<br>Spain,<br>UK | 02/01-03/<br>02 | 03/03-03/<br>17 | 03/18-04/<br>01 | 04/02-04/<br>16 | 04/17-05/<br>01 | 05/02-05/<br>16 |
| Iran | 02/20-03/<br>21 | 03/22-04/<br>05 | 04/06-04/<br>20 | 04/21-05/<br>05 | 05/06-05/<br>20 | 05/21-06/<br>04 |
| Brazil,<br>Pakistan | 02/27-03/<br>28 | 03/29-04/<br>12 | 04/13-04/<br>27 | 04/28-05/<br>12 | 05/13-05/<br>27 | 05/28-06/<br>11 |
| Nigeria | 02/28-03/<br>29 | 03/30-04/<br>13 | 04/14-04/<br>28 | 04/29-05/<br>13 | 05/14-05/<br>28 | 05/29-06/<br>12 |
| Mexico | 02/29-03/<br>30 | 03/31-04/<br>14 | 04/15-04/<br>29 | 04/30-05/<br>14 | 05/15-05/<br>29 | 05/30-06/<br>13 |
| Indonesia | 03/02-04/<br>01 | 04/02-04/<br>16 | 04/17-05/<br>01 | 05/02-05/<br>16 | 05/17-05/<br>31 | 06/01-06/<br>15 |
| Saudi<br>Arabia | 03/03-04/<br>02 | 04/03-04/<br>17 | 04/18-05/<br>02 | 05/03-05/<br>17 | 05/18-06/<br>01 | 06/02-06/<br>16 |
| Argentina | 03/04-04/<br>03 | 04/04-04/<br>18 | 04/19-05/<br>03 | 05/04-05/<br>18 | 05/19-06/<br>02 | 06/03-06/<br>17 |
| Spain | 03/06-04/<br>05 | 04/06-04/<br>20 | 04/21-05/<br>05 | 05/06-05/<br>20 | 05/21-06/<br>04 | 06/05-06/<br>19 |
| Turkey | 03/12-04/<br>11 | 04/12-04/<br>26 | 04/27-05/<br>11 | 05/12-05/<br>26 | 05/27-06/<br>10 | 06/11-06/<br>25 |

## The DEA model

In this paper, the DEA model was employed to measure the mitigation efficiency regarding the spread of COVID-19 at each stage for each country. The DEA was pioneered by Charnes *et al.* [14] based on the theoretical concept of frontier production developed by Farrell [15]. It is a linear programming technique to estimate production or cost efficiency by measuring the ratio of total inputs employed to total output produced for each decision-making unit (DMU). The mitigation of the COVID-19 transmission in each country was executed by a technology whereby *N* countries in terms of DMUs transform multiple inputs 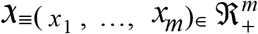 into multiple outputs 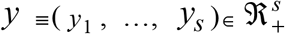. This paper employed the basic DEA model of Charnes, Coopers, and Rhodes (CCR) to calculate the mitigation efficiency of COVID-19 transmission. The CCR model, under the hypothesis of constant returns to scale, is expressed as follows:

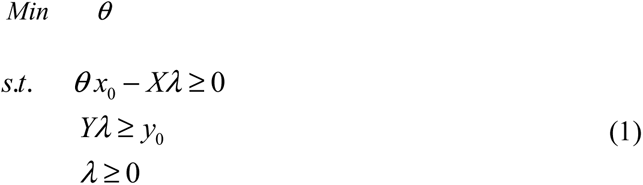

where *y*_0_ is the output, *x*_0_ is the input, *X,Y*, are the data sets in the matrices, *λ* is a semipositive vector, and *θ* represents the technical efficiency.

After the efficiency at each stage was obtained, Pearson correlation tests were conducted between the different stages at a p value < 0.01 to examine the variation in efficiency ranks across stages. The correlation tests were used to explain the impact of the efficiency ranks at previous stages on subsequent stages.

## The variables

Efficiency is described as the relative performance regarding the reduction in the COVID-19 transmission, was measured in this paper by the DEA method, and is stated in the form of an output/input ratio. The objective of the authority administration was to minimize the total confirmed cases that occurred in each stage with a given amount of resources used. Cooper *et al.* [16] suggested that the DEA technique can be easily applied to a multiple input–output framework to compare the relative efficiency among various DMUs. The information produced from the DEA analysis is valuable for identifying specific efficient units for future learning [17].

Neiderud [18] suggested that the rise of megacities may yield potential risks for new epidemics and become a threat in the world. The high human population density and close human-to-human contact are the major sources for the rapid spread of respiratory diseases or avian flu. The growth and density of the human population may work as an incubator for infectious diseases, and urbanization as a driver of disease may have a negative effect on public health (Lienhardt [19]; Hayward *et al*.[20]). Thus, the input and output variables included (1) newly confirmed cases *n*, (2) population density *d*, and (3) urbanization degree *u* in each country. As more confirmed cases represent less efficiency, the total newly confirmed cases in each stage was considered the output variable of mitigation inefficiency. High population density and urbanization degree in a country may present a greater chance of being infected; thus, population and urbanization degree are seen as input variables of mitigation inefficiency. Since mitigation efficiency is the inverse of mitigation inefficiency, newly confirmed cases *n* was treated as an output variable in this paper, and population density *d* and urbanization degree *u* were treated as output variables for the measurement of mitigation efficiency.

## Data collection

The data for accumulated confirmed cases were extracted from the daily situation reports from the WHO [1], and the total confirmed cases in each stage were calculated by the difference in the accumulated confirmed cases on the last day of each stage and the previous stage. The population density data for each country were provided by Worldometer [21], and the urbanization degree data were extracted from the World Bank [13]. The descriptive statistics for the total accumulated confirmed cases across the 6 stages (i.e., 120 days since the first confirmed case), population density and urbanization degree are presented in Table 2. By the end of Stage 6 (i.e., 120 days since the first confirmed case), the USA had 1,193,452 confirmed cases, ranking at the top of the 23 countries, while Australia had the lowest number (6,914) of confirmed cases. Korea has the highest population density at 527.30 persons per km^2,^ while Australia has a much lower population density at 3.32 persons per km^2^. Argentina has the largest urbanization degree at 92% and ranked at the top. In contrast, the urbanization degree of India is much less than the average of 71.48% based on the other countries and was only 34%.

**Table 2:** Descriptive statistics of study variables.

| Variables | Total confirmed cases $n$ | Population density $d$ (person per km<sup>2</sup>) | Urbanization degree $u$ (%) |
| --- | --- | --- | --- |
| Max. | 1,193,452 | 527.30 | 92.00 |
| Min. | 6,914 | 3.32 | 34.00 |
| Average | 190,093 | 151.40 | 71.48 |
| Standard deviation | 260,495 | 146.28 | 15.45 |

The efficiency score was calculated through the assistance of the software DEA solver 13.

## Results

The mitigation efficiency of the COVID-19 epidemic covering the first 120 days after a confirmed case for each of these countries is depicted in Fig 1. Australia and Korea rank at the top of mitigation efficiency. In contrast, the USA ranks at the bottom, followed by Brazil and Russia. The major cause affecting the efficiency ranks may be attributed to the number of total confirmed cases occurring over the whole period. The total confirmed cases in Australia and Korea in the whole period (covering 120 days since the first confirmed case) were only 6,667 cases and 10,801 cases, respectively, while the USA, Brazil, and Russia had 1,193,452, 739,503, and 272,043 cases, respectively.

**Fig. 1.**
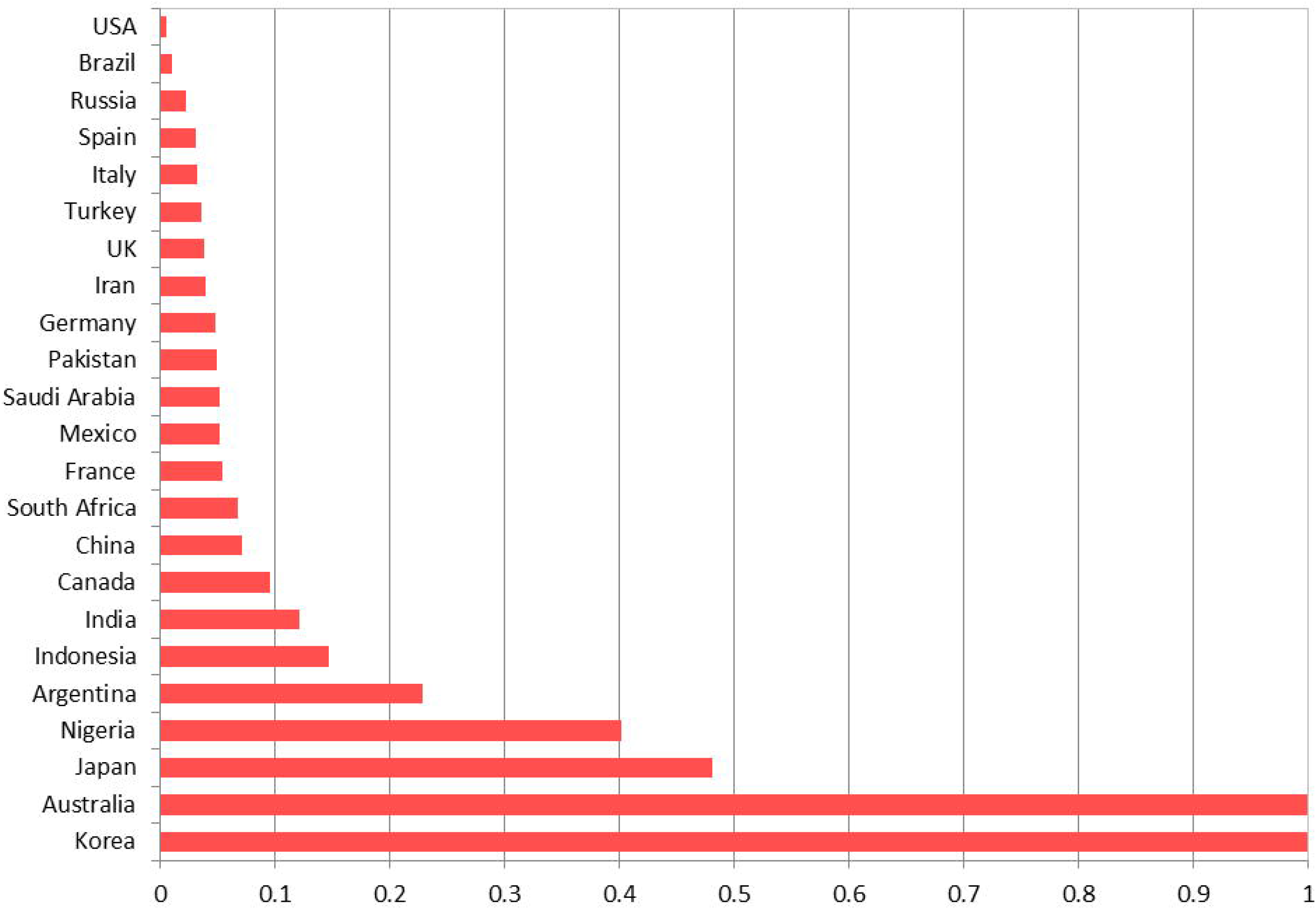
Mitigation efficiency scores among the 23 countries.

The efficiency scores and ranks at each stage for each country were also calculated according to Eq. (1). Based on the shape of the efficiency ranking trend, this paper classifies these countries into 5 types:

Type (1): An inverted U-shaped pattern including Korea, China, Italy, Spain, UK, Germany and France

This pattern in the efficiency rank trends, depicted in Fig 2, was characterized by a continual decline in mitigation efficiency from Stage 1, and after reaching the lowest point of in their efficiency ranks, these scores continued to improve until the last stage (Stage 6). Efforts to mitigate newly confirmed cases through the implementation of response strategies may have eventually achieved a certain effect stages. Passing through the peak of daily newly confirmed cases, the COVID-19 transmission was then reduced, and the efficiency started to improve through the last stage. For example, Italy ranked 14^th^ in Stage 1 and then dropped to 20^th^ 197 in Stage 2. Italy reached a peak of daily confirmed cases, amounting to 6,557 cases on 22 March 2020, which occurred in Stage 3. After Stage 3, the COVID-19 transmission in Italy improved, and the efficiency ranks rose to 13^th^ place in Stage 6. The efficiency ranks for China after Stage 3 and Korea after Stage 2 showed great improvement and attained a relatively more stable state. China ranked 21^st^ place and 22^nd^ place at Stage 1 and Stage 2, respectively, but the efficiency ranks were improved to 2^nd^ place at Stage 4 and 3^rd^ place at Stage 5 and 6 through a great number of emergency response strategies. Similar to China, Korea ranked in 10^th^ place and 13^th^ place for mitigation efficiency at Stages 1 and 2, respectively, and efficiency was improved to 6^th^ place at Stage 3 and first place at Stage 4 which was subsequently maintained to the final stage. The other countries showed similar processes, but the degree of efficiency improvement was different.

**Fig. 2.**
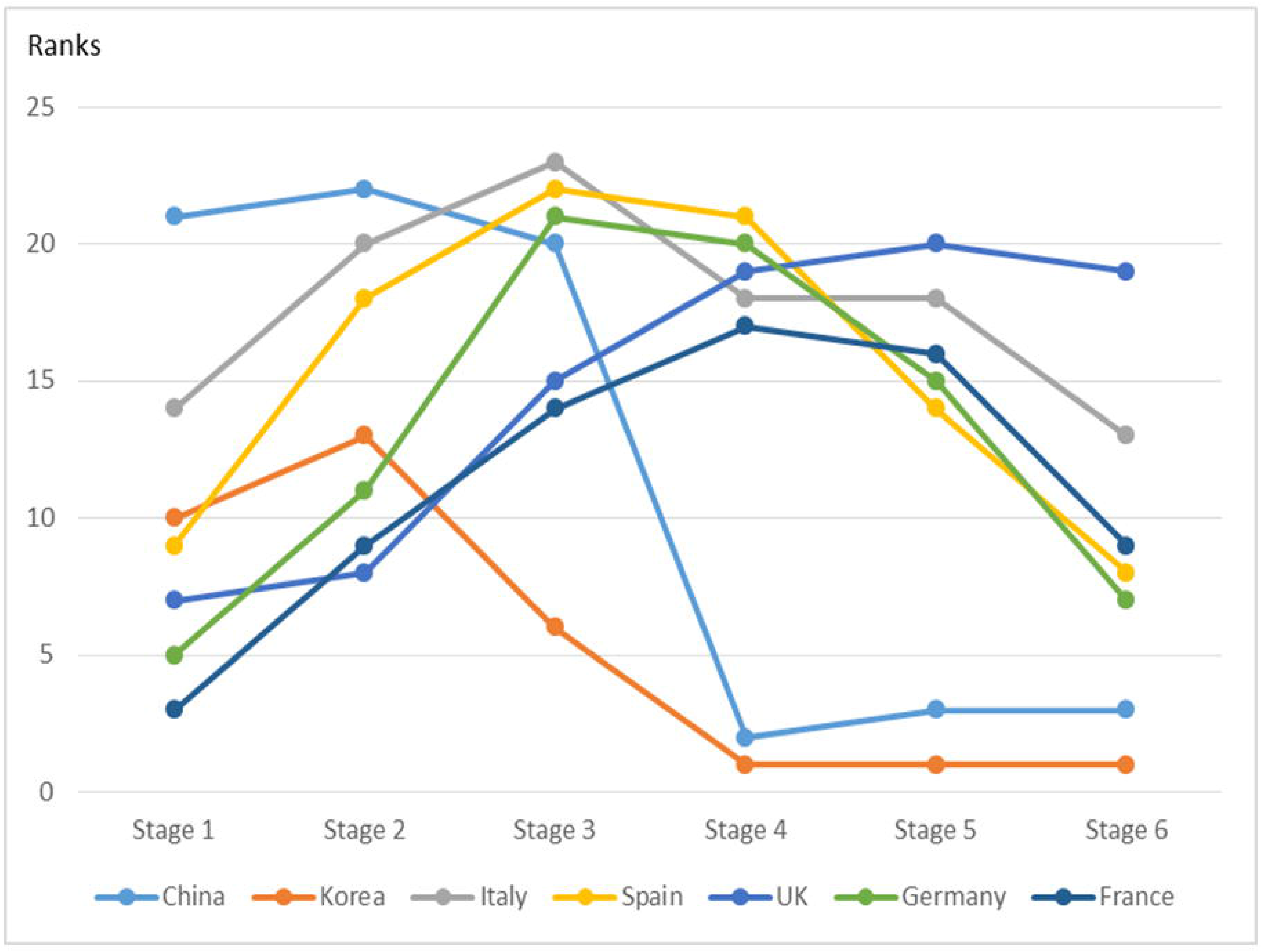
The trends in efficiency ranks for Type (1) countries.

Type (2): An inverted N-shaped pattern including Japan and Australia

The trend in efficiency ranks for Type (2) countries is depicted in Fig3. Basically, efficiency ranks fluctuated across stages, with initial improvements followed by deterioration in the middle stages, but eventually efficiency ranks improved in the final stages. For example, the efficiency ranks for Japan improved continuously from 12^th^ place in Stage 1 to 6^th^ place in Stage 2 to first place in Stage 3, then dropped to 4^th^ place in Stage 4 and 6^th^ place in Stage 5, and eventually improved to 4^th^ 219 place again.

**Fig. 3.**
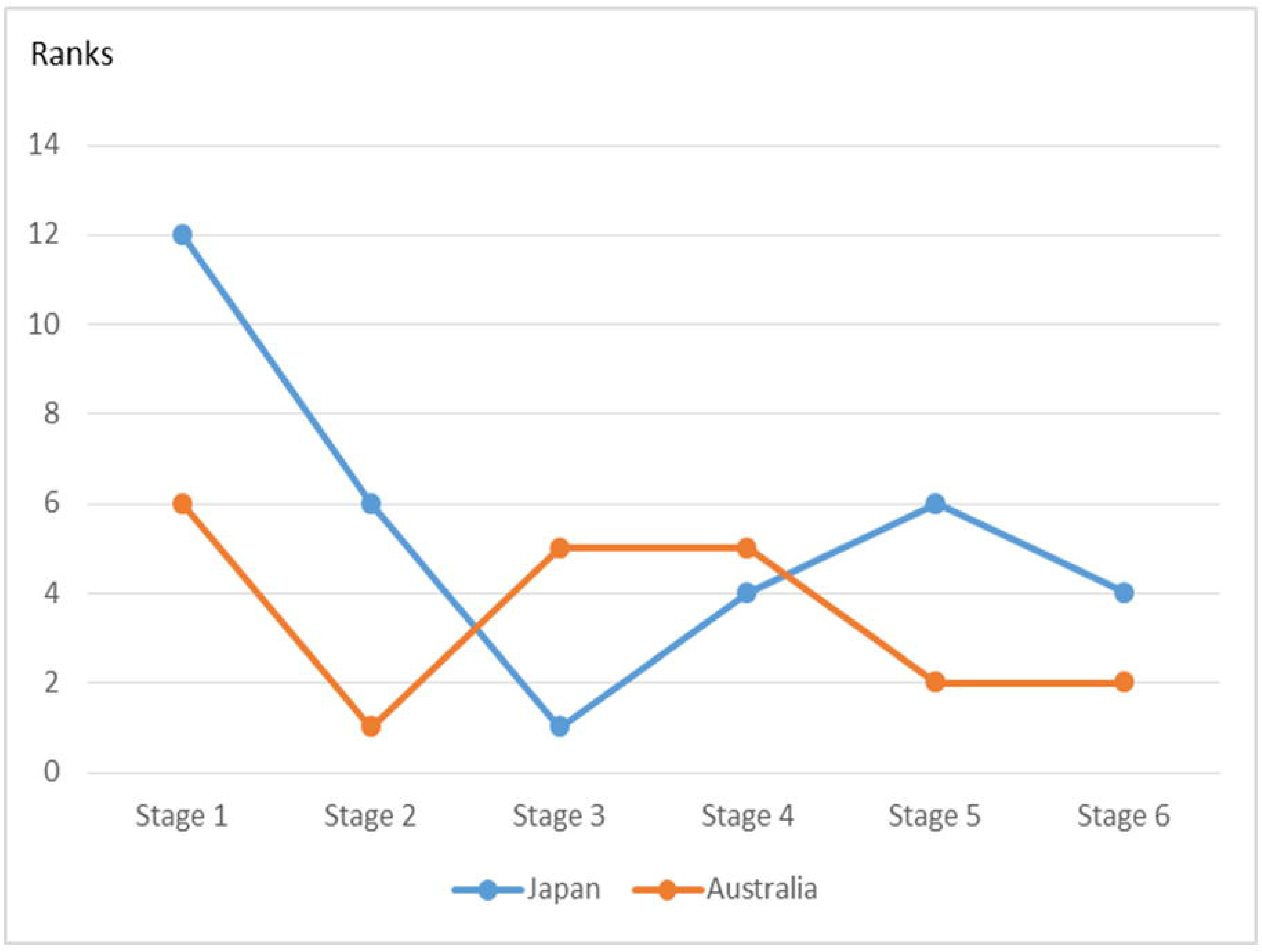
The trends in efficiency ranks for Type (2) countries.

Type (3): Continual decreases in efficiency ranks including Russia and India

The trend pattern in efficiency ranks for Type (3) countries depicted in Fig 4 is characterized by the gradual deterioration in mitigation efficiency. The efficiency ranks are not bad in the earlier stages but worsen. For example, Russia performed at the highest level regarding mitigation efficiency in Stage 1 and was ranked in first place. Unfortunately, Russia do not maintain this advantage, but its ranks continued to deteriorate to 4^th^ place in Stage 2 and finally 21^st^ place in Stage 6.

**Fig. 4.**
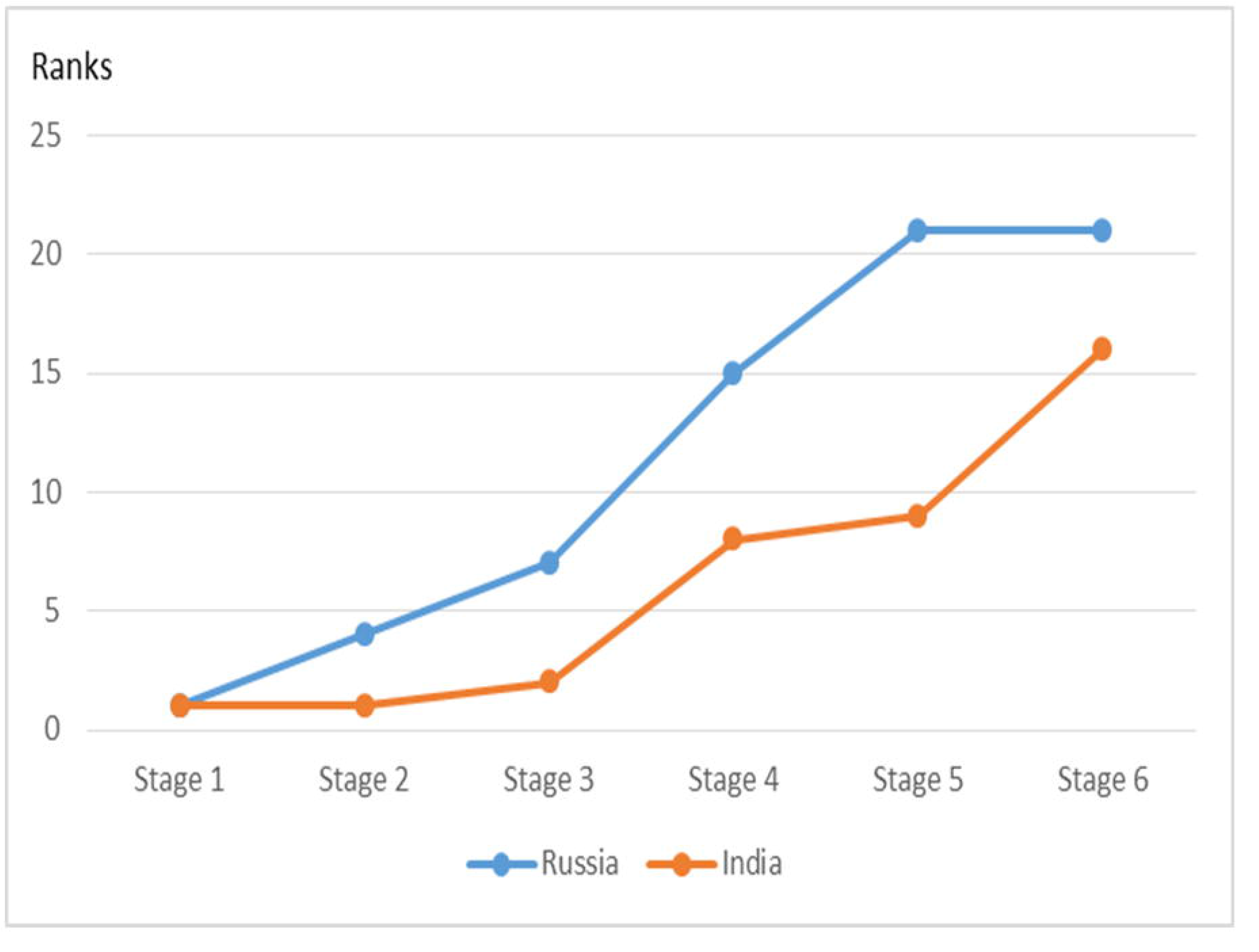
The trends in efficiency ranks for Type (3) countries.

Type (4): U-shaped pattern including the USA, Iran, Turkey, Indonesia, Pakistan, South Africa, Argentina and Brazil

This trend pattern in the efficiency ranks depicted in Fig 5 is characterized by some improvements in mitigation efficiency in the middle stages, but eventually rebounded back to a worse state. For example, the response in the USA to avoid COVID-19 transmission was not bad in Stages 1 and 2, as it ranked in 8^th^ place and 5^th^ place, respectively. However, its efficiency continually and dramatically dropped after Stage 2 and fell to 23rd place (the bottom of the ranking) in Stages 5 and 6. The efficiency improvement from Stage 1 to Stage 2 in the USA may be attributed to its prompt travel restrictions on China from 2 February 2020 and additional travel restrictions on Iran, Italy, and Korea on 29 February (Garda world, 2020). The gradual deterioration in efficiency ranking in the later stages in the USA implies that its response strategies may be ineffective for avoiding the epidemic.

**Fig. 5.**
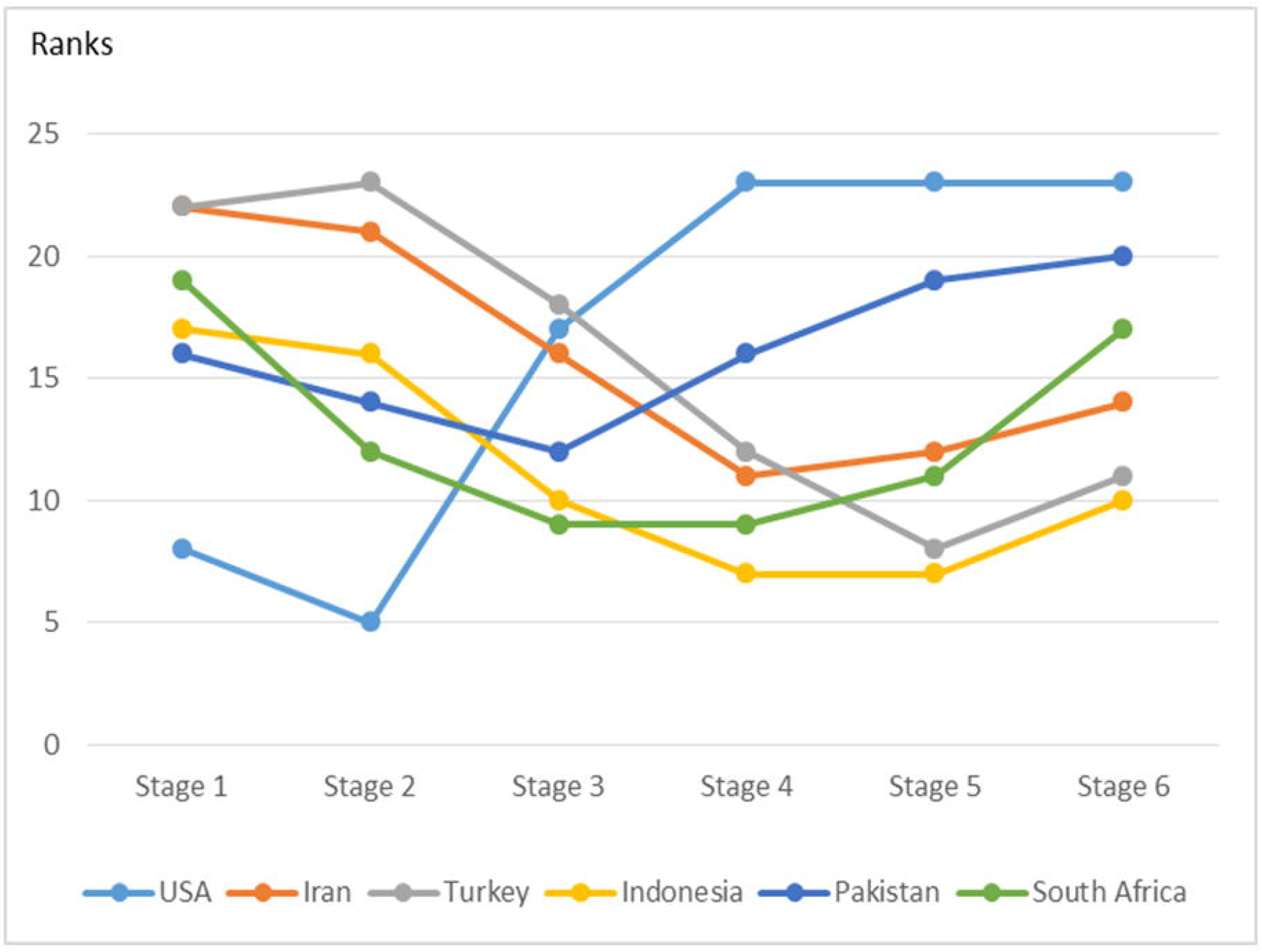
The trends in efficiency ranks for Type (4) countries.

The trend pattern in the efficiency ranking for Brazil provides a different story. From Stage 1 to Stage 6, the efficiency ranks for Brazil were not good. On 25 June 2020 (the final observation point in Stage 6) in Brazil, newly confirmed cases remained at a high level of 39,436 cases. This implies that the response strategies adopted by Brazil contained flaws.

Type (5): N-shaped and W-shaped patterns including Mexico, Nigeria and Saudi Arabia.

An N-shaped pattern for Mexico and W-shaped patterns for Nigeria and Saudi Arabia are categorized and depicted in Fig 6. At the middle stages, the efficiency ranks for these Type (5) countries fluctuated very much. For example, Mexico ranked 13^th^ place at Stage 1 and then dropped and rose in the middle stages, eventually dropping again to 17^th^ place in Stage 6. As the efficiency for these two patterns drops again in the last stages, this implies that the mitigation efficiency is not stable and that the future trends for these countries is not optimistic.

**Fig. 6.**
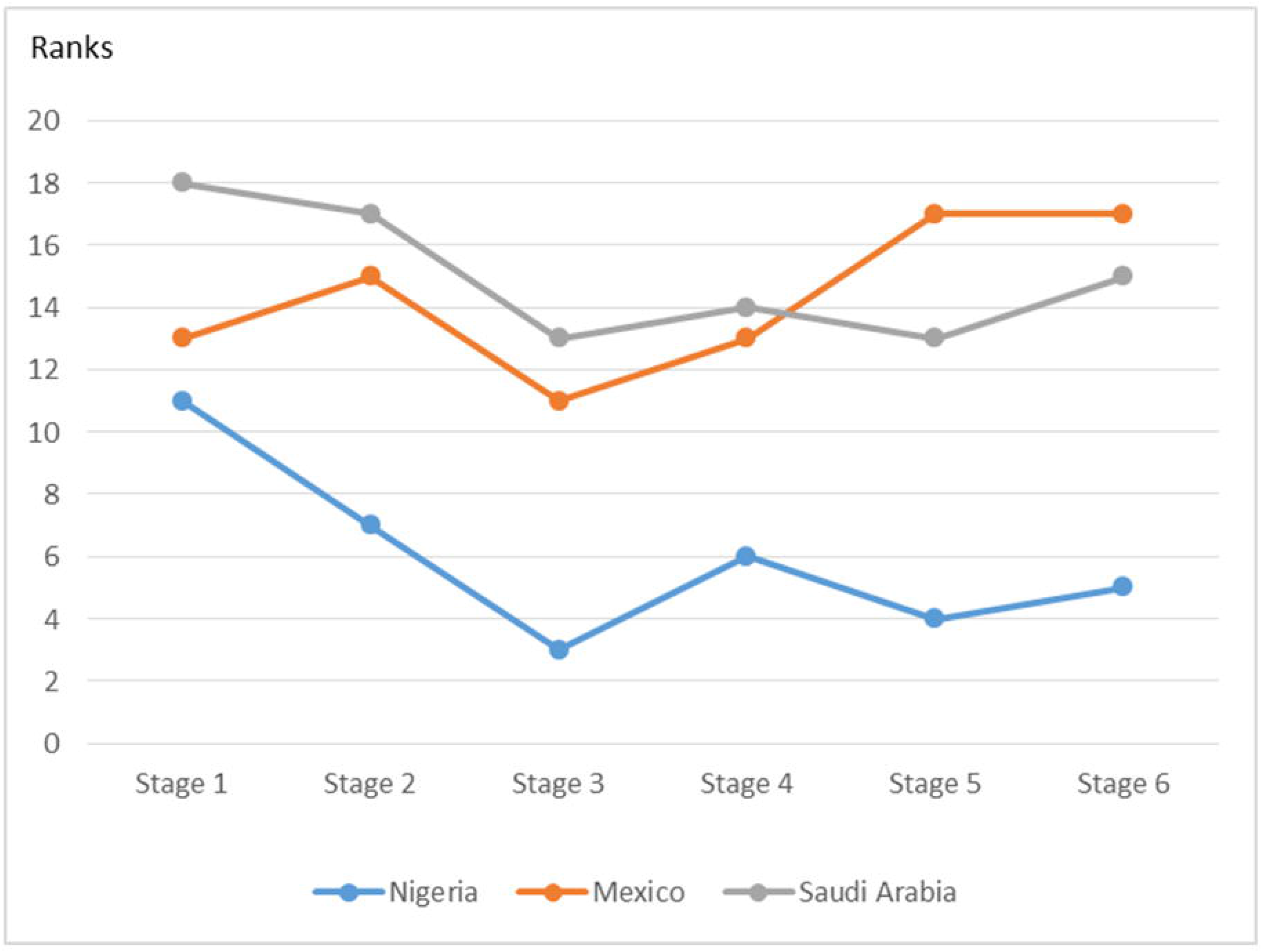
The trends in efficiency ranks for Type (5) countries.

## Discussions

The DEA analysis in this paper finds that Korea, Australia and Japan had better mitigation efficiency by 27 June 2020, while the USA, Brazil and Russia performed less efficiently and were ranked at the bottom. Michael [22] suggested that the successful experience in Korea to defeat COVID-19 may be attributed to the massive testing and effective contact tracking system. Individuals testing positive for the infection after viral tests were hospitalized at special facilities. The people who had been in contact with the infected were to remain self-quarantined for 14 days. The availability of personal protective equipment was ensured to have sufficient supply to avoid further infection at the onset of COVID-19 in Korea. In contrast, the testing capacity have not been sufficient to support the policies of a gradual reopening of the economy planned in many US states [23].

The trend patterns in efficiency ranks also revealed information about future trends regarding epidemic mitigation. Type (1) and Type (2) countries may have more optimism regarding recovery from the attack of COVID-19, as the efficiency ranks of Type (1) and Type (2) countries were good in Stage 6.

The Type (1) countries included the following 7 countries: Korea, China, Italy, Spain, the UK, Germany and France.

In addition to Korea, the other countries implemented effective responsive strategies, including extensive viral tests, lockdown, social distancing, temporary cessation of sports events, school closures, and wearing masks. In China, testing policies were promoted by expanding testing of individuals from persons with symptoms to the open public on 12 February 2020, and all levels of school were closed on 26 January 2020 [24]. China has successfully slowed the transmission of COVID-19 through a combination of lockdown, viral tests, contacting tracing and other minor strategies, including street sanitization, school closures and wearing masks. Strict lockdown and strict checks to avoid close contact between people were implemented in China after the outbreak. In less than three months, China has gradually released the strict policy of the lockdown and started to motivate the opening of economic activities. The strict lockdown, wearing masks, and social distancing implemented in China may be the major contributor to the effective prevention of transmission in a short time.

In contrast, the response of European countries such as Italy was not as prompt and urgent as that in Korea or China, and their efficiency ranks after Stage 4 were worse. For example, Italy closed their schools on 2 March 2020, asked their people to stay at home, with exceptions for daily exercise and grocery shopping, on 23 February 2020. However, the testing policy adopted in Italy focused on testing anyone with COVID-19 symptoms after 26 February [24]. However, the efficiency ranks for the UK in the later stages (Stage 4–6) were much worse than those of other European countries. In March 2020, the UK attempted to reduce the impact of COVID-19 by means of herd immunity, but later, it denied the claims of herd immunity and argued that herd immunity is a natural by-product of an epidemic [25]. Given this situation, the strategy to fight against the epidemic was delayed, and thus, the effect was reduced.

Type (2) countries consisted of only Japan and Australia with overall efficiency ranks of first and third place, respectively. In the middle stages, the efficiency ranks initially improved and then got worse. A possible cause for these changes in efficiency ranks may be explained by the low levels of viral testing in the earlier stages.

Extensive viral tests have been performed in Australia and amounted to nearly 1000 tests per 100,000 people in the population by 31 March 2020 [26]. This number continued to increase and reached 2081 tests per 100,000 people on 28 April 2020 and 3119 tests per 100,000 people on 9 May 2020 (the final observation point in Stage 6 May 2020 (the final observation point in Stage 6 for India), the viral testing rate was only 1.41 tests per 1,000 people 9 May 2020 (the final observation point in Stage 6 for Australia). The high testing rate in Australia may have been a major factor in mitigating the increase in new cases and had the best overall efficiency among these 23 countries.

In contrast, the trend in efficiency ranks for Type (3) countries showed with continual deterioration in mitigation efficiency. Compared to other countries, the coronavirus testing rate per capita in India was very low, reaching a total of 144,910 tests in a population with more than 1.3 billion people by 9 April 2020 [27]. On 14 May 2020 (the final observation point in Stage 6 for India), the viral testing rate was only 1.41 tests per 1,000 people [24]. The low testing rate may be a key factor in explaining the good performance based on the high efficiency ranking from Stage 1 to Stage 4. Without testing, no data are generated; thus, higher efficiency scores are obtained. On 27 June 2020, the total confirmed cases in India reached 508,953 cases, approximately 6.5 times the total confirmed cases in the whole period examined in this study.

At the onset of the outbreak, Russia announced temporarily banning Chinese citizens from entering Russia on 20 February 2020 [28]. This strategy may have been effective in preventing infection through imported cases from China in Stage 1 and Stage 2. Extensive testing had been conducted in Russia, including 0.32 tests per 1,000 people on 5 March 2020, 1.12 tests per 1,000 people on 22 March 2020, 4.38 tests per 1,000 people on 4 April 2020, 11.06 tests per 1,000 people on 16 April 2020, 27.04 tests per 1,000 people on 2 May 2020, and 45.61 tests per 1,000 people on 16 May 2020 (the last day of Stage 6). However, Russia’s health department admitted that the test kits were often wrong and provided false negative results. Therefore, the tested people with the virus were allowed to go home and infect other people. Thus, the real number of infected individuals was more than triple the official figure [29]. The ineffective tests may explain the continual deterioration in efficiency scores for Russia.

Type (4) countries contained the following 9 countries: the USA, Iran, Turkey, Canada, Indonesia, Pakistan, South Africa, Argentina and Brazil. Basically, if the current trends continue into the future, there is not much optimism regarding the epidemic, and these countries need to devote more effort to improving mitigation in newly confirmed cases as the efficiency ranks were bad in the final stages. Some Type(4) countries lacked testing capacity in the earlier stages of the pandemic, and thus, the amount of testing that was performed was much less than needed. Due to less viral testing than the actual need, underestimation of newly confirmed cases may have taken place and led to the illusion of efficiency improvements but eventually efficiency ranks dropped in the final stages.

In the USA, the total number of tests performed relative to the size of the population before 7 March 2020 was very low, less than 0.01 tests per 1,000 people, and the situation gradually improved in March 2020 (in Stage 3). The testing rate increased to 0.23 tests per 1,000 people by the end of March 2020 (in Stage 4) and then quickly increased to 10.43 tests per 1,000 people on 16 April 2020 (in Stage 5). On the day of the final observation point at Stage 6 (7 May 2020), the testing rate rose to 24.63 tests per 1,000 people, which seems to be a good figure compared to other countries. However, several experts have criticized that the testing levels were not sufficient to meet the need for a gradual reopening by 1 May 2020 [23]. In addition, existing flaws in other response strategies also blocked improvements in efficiency ranks for the USA. For example, the US Centers for Disease Control and Prevention (CDC) emphasized the importance of mask wearing, but Trump continued to reject being photographed in public wearing a mask [30]. Some experts have suggested that Some experts have suggested that guidelines for mask wearing have been confusing. Thus, many protesters across the country are described as people who refuse to wear a mask [31].

In fact, the USA has not been positively and seriously prepared for epidemic mitigation since the first confirmed case occurred on 23 January 2020. On 23 April 2020, Trump suggested injecting powerful disinfectant into coronavirus patients as a possible cure for COVID-19. This news resulted in criticism from many scholars and reporters and disbelief and derision worldwide [32].

The trends in efficiency ranks for Type (5) countries, including Mexico, Nigeria, and Saudi Arabia, fluctuated more than the other country types. The testing rate in Mexico ranged from 0.01 to 3.1 tests per 1,000 people during the whole period, which was much lower than that in other countries. Thus, the mitigation efficiency of Mexico ranked 17^th^ among the 23 countries in Stage 5 and Stage 6. On 13 June 2020 (the final day of Stage 6 for Nigeria), the testing rate was 0.44 tests per 1,000 people. Nigeria had a lower testing rate than Mexico, but the efficiency ranks for Nigeria were not bad. This paper reasonably suspects that the high efficiency ranks of Nigeria may be caused by an underestimation due to low viral testing rates.

The trends in efficiency ranks in Figures 2–6 also demonstrate high variation in efficiency ranking across stages for some countries. To examine the impact of efficiency ranks at the previous stage on the subsequent stage, a Pearson correlation test of efficiency scores between different stages was conducted. The results listed in Table 3 indicate that the correlation coefficient between two adjacent stages was higher and that the correlation coefficients between Stage 1 and each stage after Stage 3 were low and negative. The negative or near zero correlation coefficients between Stage 1 and Stages 4–6 implies that the efficiency ranking of the sampled countries at Stages 4–6 had been reorganized and completely differed from Stage 1. This implies that at Stage 1, some countries started to implement effective response strategies such as extensive viral testing, lockdown, wearing masks, etc. to prevent the spread of COVID-19 and thus created improved effects at Stages 4–6. In contrast, some countries purposely neglected the serious and emergent impacts arising from COVID-19 and failed to take any measures in response to the emergence of the epidemic.

**Table 3.** Correlations of mitigation efficiency between different stages.

|  | Stage 1 | Stage 2 | Stage 3 | Stage 4 | Stage 5 | Stage 6 |
| --- | --- | --- | --- | --- | --- | --- |
| Stage 1 | 1 |  |  |  |  |  |
| Stage 2 | 0.6739* | 1 |  |  |  |  |
| Stage 3 | 0.4048* | 0.5666* | 1 |  |  |  |
| Stage 4 | -0.1433* | 0.0210* | 0.4297* | 1 |  |  |
| Stage 5 | -0.0982* | 0.1918* | 0.2002* | 0.7884* | 1 |  |
| Stage 6 | -0.0824* | 0.1401* | 0.1783* | 0.7602* | 0.9828* | 1 |

\* Marks statistical significance at the 95% confidence level

In contrast, the correlation coefficient was 0.788 between Stage 4 and Stage 5, 0.760 between Stage 4 and Stage 6, and 0.983 between Stage 5 and Stage 6. These high correlation coefficients imply that the relative efficiency ranks among these countries became stable because their response strategies had stabilized.

The efficiency ranks in some countries showed a high degree of fluctuation across stages, especially the Type (5) countries. The high fluctuation in efficiency ranks implied that good efficiency rankings at a particular stage were only temporary and may have deteriorated in the next stage. The mitigation efficiency rankings for Type (3) countries continually worsened from Stage 1 to Stage 6. Thus, the Type (3) countries cannot recover from the attack of COVID-19 in a short time and have to adopt stricter response policies to mitigate the spread of COVID-19. Type (4) countries executed a U-shaped pattern, demonstrating temporarily improved ranks in the middle stages but eventually the ranking turned bad in the final stages.

Both an inverted U-shaped (Type 1) and an inverted N-shaped (Type 2) pattern in the trends in efficiency ranks seemed to be a good sign of improvement as the efficiency ranks increased in the last stages. The probability of recovering from the attack of COVID-19 for Type (1) and (2) patterns is higher than other patterns.

Nevertheless, the overall efficiency was calculated based on the whole period covering 120 days since the first confirmed case. The efficiency obtained was only temporary and may change for the better or worse if the assessment stage was extended to cover more days.

In the beginning of June 2020, the infectious disease COVID-19 remained a high risk in the world, but many countries have attempted to lift the state of lockdown, restart the economy and take actions, as their governments have considered that the number of confirmed cases was greatly reduced and that newly diagnosed cases may be considered sporadic cases. For example, Trump attempted to end the lockdown and the stay-at-home order and to reopen schools at the beginning of June 2020.

There was a high correlation between efficiency scores in two adjacent stages, but it was still difficult to predict the epidemic stability of the next stage based on the previous stage. Thus, the data of the newly confirmed cases for the current dates are only for reference to determine the timing of restarting the economy.

This paper suggests that an epidemic stability indicator in combination with a trend pattern of efficiency ranks such as Type (1) or (2) may be employed to judge the appropriateness of any measures to lift the response strategies such as travel restrictions, stay-at-home orders, and mask wearing. In this paper, epidemic stability *(ES)* is defined as the recovery status from the epidemic, and the indicator *ES* is presented by measuring an average increase in proportion of confirmed cases to population (PCCP) during the period of the last day of Stage 6 and a day designated to restart the economies, expressed as follows:

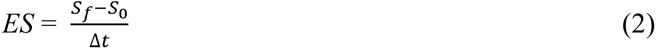

where *s_f_* and *s_o_* denote the PCCP on the last day of Stage 6 and the designated day, respectively, and *Δt* represents the period between the two dates.

A numerical example is presented in this paper. As an example, if it was proposed that the travel restrictions are lifted on the designed date of 27 June 2020, *ES*, *s_f_*, *s_o_* and *Δt* were calculated according to Eq. (2) for these 23 countries, and the result is listed in Table 4, where *s_f_* and *s_o_* are measured by cases per 100,000 persons, *Δt* in days, and *ES* by cases per 1,000,000 persons.

**Table 4.** The epidemic stability for each country by ranks.

| DMU | $S_0$ | $S_f$ | $\Delta t$ | $ES$ | Ranks |
| --- | --- | --- | --- | --- | --- |
| China | 5.81 | 5.92 | 74 | 0.01 | 1 |
| Japan | 12.04 | 14.47 | 59 | 0.46 | 2 |
| Korea | 21.07 | 24.68 | 54 | 0.68 | 3 |
| Australia | 27.11 | 29.78 | 30 | 0.89 | 4 |
| Nigeria | 7.06 | 11.3 | 15 | 2.83 | 5 |
| Indonesia | 13.99 | 18.74 | 12 | 3.95 | 6 |
| Germany | 203.51 | 230.64 | 46 | 5.90 | 7 |
| Italy | 368.99 | 396.88 | 43 | 6.49 | 8 |
| India | 5.65 | 36.88 | 44 | 7.10 | 9 |
| Spain | 492.32 | 530.22 | 42 | 9.02 | 10 |
| France | 209.24 | 239.23 | 30 | 10.00 | 11 |
| Turkey | 227.25 | 230.63 | 2 | 16.92 | 12 |
| Canada | 180.16 | 271.9 | 47 | 19.52 | 13 |
| Pakistan | 54.12 | 90.04 | 16 | 22.45 | 14 |
| UK | 348.69 | 455.71 | 42 | 25.48 | 15 |
| Iran | 191.32 | 259.22 | 21 | 32.33 | 16 |
| Mexico | 103.91 | 157.41 | 14 | 38.21 | 17 |
| Argentina | 72.54 | 116.07 | 10 | 43.53 | 18 |
| Russia | 186.41 | 430.09 | 42 | 58.02 | 19 |
| USA | 360.56 | 727.36 | 51 | 71.92 | 20 |
| South Africa | 141.45 | 210.07 | 8 | 85.78 | 21 |
| Saudi Arabia | 379.3 | 501.46 | 13 | 93.97 | 22 |
| Brazil | 347.9 | 577.77 | 16 | 143.67 | 23 |

Table 4 indicates that India has the lowest value of *s_o_* (PCCP in 120 days). amounting to 5.65 cases per 100,000 persons, and slightly lower value than China(5.81 cases per 100,000 persons). In contrast, Spain and Saudi Arabia have the highest value of *s_o_*, amounting to 492.32 and 379.30 cases per 100,000 persons, respectively, which is much higher than the average of 172.19 cases per 100,000 persons. However, the ranking of the PCCP on 27 June 2020 (*s_f_*) changes very much. China ranks at the 8 top with the lowest *s_f_*, amounting to 5.92 cases per 100,000 persons. The PCCP in India increases very much from 5.65 at *s_o_* 36.88 cases per million at *s_f_*. The USA has the highest value at *s_f_*, amounting to 727.37 cases per 100,000 persons.

Table 4 also demonstrates that the *ES* in China, Japan, Korea and Australia is much better than that in the other countries, amounting to 0.01, 0.46, 0.68, and 0.89 cases per million persons per day during the period between the last day of Stage 6 and 27 June 2020. In contrast, the *ES* in Brazil, Saudi Arabia, South Africa and the USA reaches 143.67, 93.97, 85.78 and 71.92 cases per million persons per day, respectively. Based on the value of *ES*, it is suggested that the future trends regarding the pandemic in Brazil, Saudi Arabia, South Africa and the USA are not optimistic, and it is full of challenges.

Low values of epidemic stability imply that the trend regarding the epidemic has attained a stable state and approached zero confirmed cases. Thus, China, Japan, Korea and Australia seem to have recovered from the attack of COVID-19, while Brazil, Saudi Arabia, South Africa and the USA remain engaged in the battle fighting against COVID-19 and are required to devote more effort to create new opportunities. On 27 June 2020, China, Japan, Korea, and Australia had 24, 100, 51, 37 daily newly confirmed cases (WHO, 2020), much lower than the peak of newly confirmed cases for each country. In contrast, at the end of June 2020, Brazil and the USA have continually set new records for daily, newly confirmed cases. The number of newly confirmed cases on 27 June 2020 was 39,483, 3,938, 6,215, and 40,526 cases for Brazil, Saudi Arabia, South Africa and the USA, respectively [1].

On 30 June 2020, the European Council announced the release of travel restrictions from 1 July 2020 for residents of recommended countries, including Australia, Japan, Korea, China and Canada [33]. As indicated in Table 4, China, Japan, Korea and Australia ranked first to fourth in epidemic stability. Canada is slightly behind in 13^th^ place. To examine the appropriateness of lifting the travel restrictions at the external borders for residents of these countries, this paper uses the data for 27 June 2020 as an example. On that day, the number of newly confirmed cases in China, Japan, Korea, Australia and Canada was 24, 100, 51, 37, and 380, respectively, equivalent to the stability of 0.0168, 0.791, 0.995, 1.451 and 10.068 cases per million per day. The *ES* on 27 June 2020 in China, Japan, Korea and Australia was much lower than the value of Germany’s *ES* (Table 4). This implies that the spread of COVID-19 has been controlled in these countries and is more stable than in Germany. The *ES* value on 27 June 2020 for Canada was nearly the same as that for France, as indicated in Table 4. However, Canada has executed a U-shaped pattern for the trend in efficiency ranks, and this paper suggests that the EU wait and observe the efficiency trend and the newly confirmed cases for Canada. Thus, this paper suggests that the lifting of travel restriction for these countries, with the exception of Canada, is quite reasonable based on the indicator of epidemic stability and the trends in efficiency ranking presented in this paper.

## Conclusions

At the onset of COVID 19 infection in a population, a massive testing program and effective tracing system on infected people were implemented in some countries, such as China and Korea. Based on the trends in efficiency ranks and the epidemic stability indicators, China, Korea, Japan and Australia have performed better than other counties. Thus, this paper suggests massive testing together with other strategies, such as contact tracing, lockdown, mask wearing, and social distancing is significantly effective in mitigating the transmission of COVID-19. Testing suspected persons identified through contact tracing and reducing interpersonal contacts through complete or partial lockdown also play important roles in reducing the number of confirmed cases. Castillo *et al*. [34] examined the effect of the stay-at-home policy on COVID-19 infection rates and found that the infection rate decreased from 0.113/day pre policy to 0.047/day post policy. Ferguson *et al*. [35] found that a lockdown may result in an average reduction in COVID-19 transmission by 50%, school closure by 20%, and other measures by approximately 10% (cited from Willis *et al*. [36]). Some other studies have also presented the same conclusions that nonpharmaceutical interventions may effectively prevent the spread of infection [37, 38].

Pearson’s correlation tests were also performed in this paper to examine the impact of efficiency at earlier stages on subsequent stages and found that efficiency ranks for each country dramatically changed across stages. Thus, the data of newly confirmed cases occurring at the present time are only for reference. The major contribution of this paper was to develop an epidemic stability measure integrating the trends in efficiency ranks to judge the appropriateness to reopen economies. Having not reached an appropriate level of epidemic stability, economic reopening may damage the anti-epidemic achievements from the earlier stages and lead to a second wave of epidemic with exponential growth in the number of newly confirmed cases.

## Data Availability

Data for accumulated confirmed cases were extracted from the daily Coronavirus disease (COVID-2019) situation reports from the WHO, Available from: https://www.who.int/emergencies/diseases/novel-coronavirus-2019/situation-reports.

